# Genome-Wide Association Identifies the First Risk Loci for Psychosis in Alzheimer Disease

**DOI:** 10.1101/2020.08.07.20139261

**Authors:** Mary Ann A. DeMichele-Sweet, Lambertus Klei, Byron Creese, Janet C. Harwood, Elise A. Weamer, Lora McClain, Rebecca Sims, Isabel Hernandez, Sonia Moreno-Grau, Lluís Tárraga, Mercè Boada, Emilio Alarcón-Martín, Sergi Valero, NIA-LOAD Family Based Study Consortium, Alzheimer’s Disease Genetics Consortium (ADGC), Yushi Liu, Basavaraj Hooli, Dag Aarsland, Geir Selbaek, Sverre Bergh, Arvid Rongve, Ingvild Saltvedt, Håvard K. Skjellegrind, Bo Engdahl, Eystein Stordal, Ole A. Andreassen, Srdjan Djurovic, Lavinia Athanasiu, Davide Seripa, Barbara Borroni, Diego Albani, Gianluigi Forloni, Patrizia Mecocci, Alessandro Serretti, Diana De Ronchi, Antonis Politis, AddNeuroMed Consortium, Julie Williams, Richard Mayeux, Tatiana Foroud, Agustin Ruiz, Clive Ballard, Peter Holmans, Oscar L. Lopez, M. Ilyas Kamboh, Bernie Devlin, Robert A. Sweet

**Affiliations:** Departments of Psychiatry, University of Pittsburgh, Pittsburgh, PA; Departments of Neurology, University of Pittsburgh, Pittsburgh, PA; Departments of Human Genetics, University of Pittsburgh, Pittsburgh, PA; University of Exeter Medical School, College of Medicine and Health, Exeter, UK; Norwegian, Exeter and King’s College Consortium for Genetics of Neuropsychiatric Symptoms in Dementia; Division of Psychological Medicine and Clinical Neuroscience, School of Medicine, Cardiff University, Cardiff, UK; Research Center and Memory Clinic Fundació ACE, Institut Català de Neurociències Aplicades. Universitat Internacional de Catalunya, Barcelona, Spain; CIBERNED, Network Center for Biomedical Research in Neurodegenerative Diseases, National Institute of Health Carlos III, Spain; See Supplementary Acknowledgments for the contributors and their affiliations; Global Statistical Science, Lilly Research Laboratories, Eli Lilly and Company, Indianapolis, IN; Neurodegeneration Research, Lilly Research Laboratories, Eli Lilly and Company, Indianapolis, IN; Department of Old Age Psychiatry, Institute of Psychiatry, Psychology and Neuroscience, King’s College London and Centre for Age-Related Medicine, Stavanger University Hospital; Norwegian National Advisory Unit in Ageing and Health, Vestfold Hospital Trust, Tønsberg, Norway and Department Geriatric Medicine, Oslo University Hospital, Oslo, Norway, and Faculty of Medicine, University of Oslo, Oslo, Norway; Research Centre of Age-related Functional Decline and Disease, Innlandet Hospital Trust, Pb 68, Ottestad 2312, Norway; Department of Research and Innovation, Helse Fonna, Haugesund and Department of Clinical Medicine (K1), University of Bergen, Bergen, Norway; Geriatric Department, St. Olav Hospital, University Hospital of Trondheim, Norway and Department of Neuromedicine and Movement science, Norwegian University of Science and Technology (NTNU), Norway; HUNT Research Centre, Department of Public Health and Nursing, Faculty of Medicine and Health Sciences, Norwegian University of Science and Technology (NTNU), Levanger, Norway and Levanger Hospital, Nord-Trøndelag Hospital Trust, Levanger, Norway; Norwegian Institute of Public Health, Oslo, Norway; Department of Mental Health, Norwegian University of Science and Technology, Trondheim, 43 Norway; Department of Medical Genetics, Oslo University Hospital, Oslo, Norway and NORMENT, Department of Clinical Science, University of Bergen, Bergen, Norway; NORMENT, Institute of Clinical Medicine, University of Oslo, Oslo, Norway; Department of Hematology and Stem Cell Transplant, Vito Fazzi Hospital, Lecce, Italy; Centre for Neurodegenerative Disorders, Department of Clinical and Experimental Sciences, University of Brescia, Italy; Neuroscience Department, Istituto di Ricerche Farmacologiche Mario Negri IRCCS, Milan, Italy; Institute of Gerontology and Geriatrics, Department of Medicine and Surgery, University of Perugia, Italy; Department of Biomedical and NeuroMotor Sciences, University of Bologna, Bologna, Italy; 1st Department of Psychiatry, Eginition Hospital, Medical School, National & Kapodistrian University of Athens, Athens, Greece; UK Dementia Research Institute @ Cardiff, School of Medicine, Cardiff University, Cardiff, UK; Departments of Neurology, Psychiatry and Epidemiology, Columbia University, New York, NY; Medical and Molecular Genetics, Indiana University School of Medicine, Indianapolis, IN; University of Exeter Medical School, Exeter, UK; Norwegian, Exeter and King’s College Consortium for Genetics of Neuropsychiatric Symptoms in Dementia; NORMENT Centre, Institute of Clinical Medicine, University of Oslo, and Oslo University Hospital, Oslo, Norway; MRC Centre for Neuropsychiatric Genetics and Genomics, Division of Psychological Medicine and Clinical Neurosciences, Cardiff University, Cardiff, UK

## Abstract

Psychotic symptoms, defined as the occurrence of delusions or hallucinations, are frequent in Alzheimer disease (AD with psychosis, AD+P). AD+P affects ∼50% of individuals with AD, identifies a subgroup with poor outcomes, and is associated with a greater degree of cognitive impairment and depressive symptoms, compared to subjects without psychosis (AD-P). Although the estimated heritability of AD+P is 61%, genetic sources of risk are unknown. We report a genome-wide meta-analysis of 12,317 AD subjects, 5,445 AD+P. Results showed common genetic variation accounted for a significant portion of heritability. Two loci, one in *ENPP6* (rs9994623, O.R. (95%CI) 1.16 (1.10, 1.22), p=1.26×10^−8^) and one spanning the 3’-UTR of an alternatively spliced transcript of *SUMF1* (rs201109606, O.R. 0.65 (0.56-0.76), p=3.24×10^−8^), had genome-wide significant associations with AD+P. Gene-based analysis identified a significant association with *APOE*, due to the *APOE* risk haplotype ε4. AD+P demonstrated negative genetic correlations with cognitive and educational attainment and positive genetic correlation with depressive symptoms. We previously observed a negative genetic correlation with schizophrenia; instead, we now found a stronger negative correlation with the related phenotype of bipolar disorder. Analysis of polygenic risk scores supported this genetic correlation and documented a positive genetic correlation with risk variation for AD, beyond the effect of ε4. We also document a small set of SNPs likely to affect risk for AD+P and AD or schizophrenia. These findings provide the first unbiased identification of the association of psychosis in AD with common genetic variation and provide insights into its genetic architecture.

## Introduction

Psychotic symptoms, defined as the occurrence of delusions or hallucinations, constitute a phenotype within Alzheimer disease (AD+Psychosis, AD+P) that affects ∼ 40% to 60% of individuals with AD and is associated with poor outcomes.^1^ In comparison to AD subjects without psychosis (AD-P), AD+P subjects have greater cognitive impairments and experience more rapid declines in cognition and function that begin prior to psychosis onset.^2-9^ AD+P is also often associated with increased rates of concurrent neuropsychiatric symptoms, including agitation,^10^ aggression,^11,12^ and depression.^5,13-15^ As a consequence, AD+P is associated with increased rates of other poor outcomes, including greater distress for family and caregivers,^16^ higher institutionalization rates,^17-20^ worse health,^21^ and increased mortality^22^ compared to AD-P patients.

The AD+P phenotype is well suited for genetic studies when careful attention is paid to excluding potential phenocopies of both AD+P and AD-P. For example, we have shown that the heritability of AD+P is greatest when requiring the presence of multiple or recurrent psychotic symptoms, rather than a one-time occurrence of a single symptom.^23^ Similarly, because psychotic symptoms typically emerge in the transition from mild to moderate stages of AD,^5^ individuals without psychosis who are still in the early stages of disease may later manifest psychosis, and therefore, need to be excluded from analysis. Using these approaches to phenotypic characterization, we have previously reported familial aggregation of AD+P,^24^ which has since been replicated in two independent cohorts.^5,25^ We further estimated the heritability of the presence or absence of psychosis in AD at 61%.^23,26^

Thus, AD+P is likely to be strongly influenced by genetic variation. To date, no study has identified genome-wide significant associations with AD+P, largely due to the small sample sizes of prior studies. However, in prior reports we identified negative genetic correlation of AD+P risk with risk for schizophrenia.^27,28^ We now report a large genome-wide association meta-analysis of 12,317 AD subjects with and without psychosis. We identified two loci with genome-wide significant associations with AD+P, in *ENPP6* and *SUMF1*. In gene-based analyses, only *APOE* (p=1.23×10^−6^) reached the criterion for genome-wide significance. AD+P was negatively genetically correlated with educational attainment and positively with depressive symptoms. Surprisingly, AD+P was not significantly genetically correlated with schizophrenia, but it was negatively correlated with bipolar disorder. Analysis of polygenic risk scores derived from schizophrenia (PRS_SZ_), and bipolar disorder (PRS_BP_) GWAS, support these genetic correlations. However, the relationship of schizophrenia risk to AD+P appears subtle, with some established risk SNPs for schizophrenia likely to confer risk for AD+P, while others confer protection.

## Materials and Methods

### Subjects

This study analyzed samples from 12,317 subjects diagnosed with possible, probable,^29^ and when available, autopsy-confirmed definite^30^ Alzheimer disease (for subject characteristics see **Table 1A**). Diagnoses were made based on diagnostic evaluations, cognitive testing, and in some cases neuropathologic assessment, conducted during subjects’ participation in the following eight source programs as previously described: the Fundació ACE Barcelona Alzheimer Treatment and Research Center (ACE/GR@ACE),^31-33^ a Consortium of National Institute on Aging Alzheimer Disease Centers (ADC),^34^ Eli Lilly and Company (LILLY),^35,36^ the Norwegian, Exeter and King’s College Consortium for Genetics of Neuropsychiatric Symptoms in Dementia (NEXGENS),^37-42^ the National Institute on Aging’s Late Onset Alzheimer’s Disease Family Study (NIA-LOAD),^5,26^ the National Institute of Mental Health Genetics Initiative AD Cohort (NIMH),^24^ the University of Pittsburgh Alzheimer Disease Research Center (PITT ADRC),^43,44^ and the MRC genetic resource for Late-onset AD included in the Genetic and Environmental Risk in AD Consortium (UK-Cardiff).^27,31,45^ Collection of clinical data and genetic samples were approved by each source program’s local Institutional Review Board or Medical Ethics Committee, as appropriate.

**Table 1A.** Subject Characteristics.

|  | **AD-P**<br>6,872 (55.8%)<br>**N (%) or Mean (SD)** | **AD+P**<br>5,445 (44.2%)<br>**N (%) or Mean (SD)** | **Total**<br>12,317 (100.0%)<br>**N (%) or Mean (SD)** |
| --- | --- | --- | --- |
| **Female** | 4,008 (58.3) | 3,649 (67.0) | 7,657 (62.2) |
| **Age of Onset<sup>1</sup>** | 74 (8.3) | 73.4 (8.0) | 73.8 (8.2) |
| **Age at Consent** | 75.6 (7.0) | 77.0 (6.9) | 76.2 (7.0) |
| **Age at Last Visit<sup>1</sup>** | 80.5 (8.1) | 81.3 (7.7) | 80.9 (7.9) |
| **Last MMSE<sup>1</sup>** | 16.0 (6.5) | 13.7 (6.8) | 15.0 (6.8) |
| **Last CDR<sup>1</sup>** |  |  |  |
| 0.0 | 2 (0.0) | 5 (0.1) | 7 (0.1) |
| 0.5 | 244 (3.8) | 186 (3.8) | 430 (3.8) |
| 1.0 | 2,469 (38.4) | 967 (19.6) | 3,436 (30.2) |
| 2.0 | 1,560 (24.3) | 1,586 (32.2) | 3,146 (27.7) |
| 3.0 | 986 (15.3) | 1,340 (27.2) | 2,326 (20.5) |
| 4.0 | 759 (11.8) | 482 (9.8) | 1,241 (10.9) |
| 5.0 | 410 (6.4) | 363 (7.4) | 773 (6.8) |

AD-P: Alzheimer disease without psychosis; AD+P: Alzheimer disease with psychosis; MMSE:
Mini mental state exam; CDR: CDR® Dementia Staging Instrument; <sup>1</sup>Data not available for some subjects/source programs; see **Supplementary Tables S8-S15** for details.

### Characterization of Psychosis

Subjects were characterized for the presence or absence of delusions and hallucinations within the individual source programs (including their sub-studies) using the CERAD behavioral rating scale^46^ (PITT ADRC and NIA-LOAD), Neuropsychiatric Inventory Questionnaire (NPI-Q,^47^ NIA-LOAD, ADC, NEXGENS), NPI-Q Spanish Language Version^48^ (ACE/GR@ACE), NPI^49^ (UK-Cardiff, NEXGENS, LILLY), and Brief Psychiatric Rating Scale^50^ (NIMH). Each of these instruments has established reliability in AD,^5,51^ and we have previously used all successfully in analyses of psychosis in AD subjects.^4,5,7,23,43^ AD+P was defined by the presence of persistent hallucinations or delusions throughout the course of dementia, AD-P was defined by the absence of all symptoms at all assessments. However, because psychotic symptoms typically emerge in the transition from mild to moderate stages of AD^5^, individuals without psychosis, but who were still in the early stages of disease at their last assessment (CDR® Dementia Staging Instrument^52^ score < 1, mini-mental state examination score^53^ > 20), were considered to be at substantial risk of developing AD+P later in their course. Thus, these individuals were excluded from the analysis. We have used these approaches to characterizing and defining AD+P and AD-P in multiple studies demonstrating the heritability and association with genetic variation of the AD+P phenotype.^5,23,24,26-28,54^

For additional detail of each source program’s clinical assessment methodology and demographics, see **Supplementary Material**.

### Genotypes

Six of the eight program sources provided us with either blood (ACE/GR@ACE) or DNA samples (PITT ADRC, UK-Cardiff, NIA-LOAD, ADC, NIMH), all of which were processed by the Genomics Core Lab at the University of Pittsburgh. Genomic DNA was extracted from whole blood samples using the Qiamp Blood Mini kit (Qiagen, Valencia, CA). All DNA was quantitated by Pico Green (Thermo Fisher, Pittsburgh, PA) and diluted to a DNA concentration of 23ng/µl. Samples were genotyped at the Children’s Hospital of Philadelphia (CHoP, Philadelphia, PA) using Illumina’s Global Screening Array (Illumina, San Diego, CA). Prior to genotyping, ChoP confirmed DNA concentrations by Pico Green assay, and performed WGA on samples when necessary.

In addition to the above-mentioned blood and DNA samples, ACE/GR@ACE, LILLY, and NIA-LOAD provided us with single nucleotide polymorphism (SNP) array data. For the ADC, SNP array data was provided by the Alzheimer’s Disease Genetics Consortium (ADGC). NEXGENS provided genome-wide association (GWA) statistics for the comparison of AD-P and AD+P. Additional details of the generation of SNP array data for all programs can be found in the **Supplementary Material**.

### Analysis

Data from the eight program sources were processed as four cohorts (Phase 1, Phase 2, GR@ACE, and NEXGENS), based on timing of receipt of the data. Data processing, QC, and statistical analyses were uniform across three of the cohorts for which there were genotypes (Phase 1, Phase 2, GR@ACE), whereas only summary statistics were available for the fourth cohort (NEXGENS). All cohorts were analyzed separately for GWA, then statistics per SNP from these analyses were combined by meta-analysis using METAL.^55^ Below we describe quality control procedures for the three genotyped cohorts. For more detail of other methods see the **Supplementary Material**. Additional details for the NEXGENS cohort have been described previously.^56^ Methods were implemented within Plink^57,58^ unless otherwise noted.

Quality Control (QC) was completed by both genotype and by sample from Phase 1, Phase 2, and GR@ACE. After QC, 6,872 AD-P and 5,445 AD+P subjects, distributed across the four cohorts, remained for analysis (**Table 1B**). We determined ancestry using GemTools analysis^59^ of a subset of autosomal SNPs with non-call rate < 0.001 and MAF > 0.05 for Phase 1, Phase 2, and GR@ACE. These SNPs were pruned such that, within a 50 SNP block and a 5 SNP step-size, the linkage disequilibrium r^2^ < 0.01. For each of the three cohorts, a different subset of SNPs was chosen for ancestry analysis, and the resulting ancestry plots were used to identify the samples in the major European ancestry cluster. Analysis of NEXGENS^56^ was restricted to individuals of European ancestry using genetic principal components computed by EIGENSTRAT.^60^

**Table 1B.**
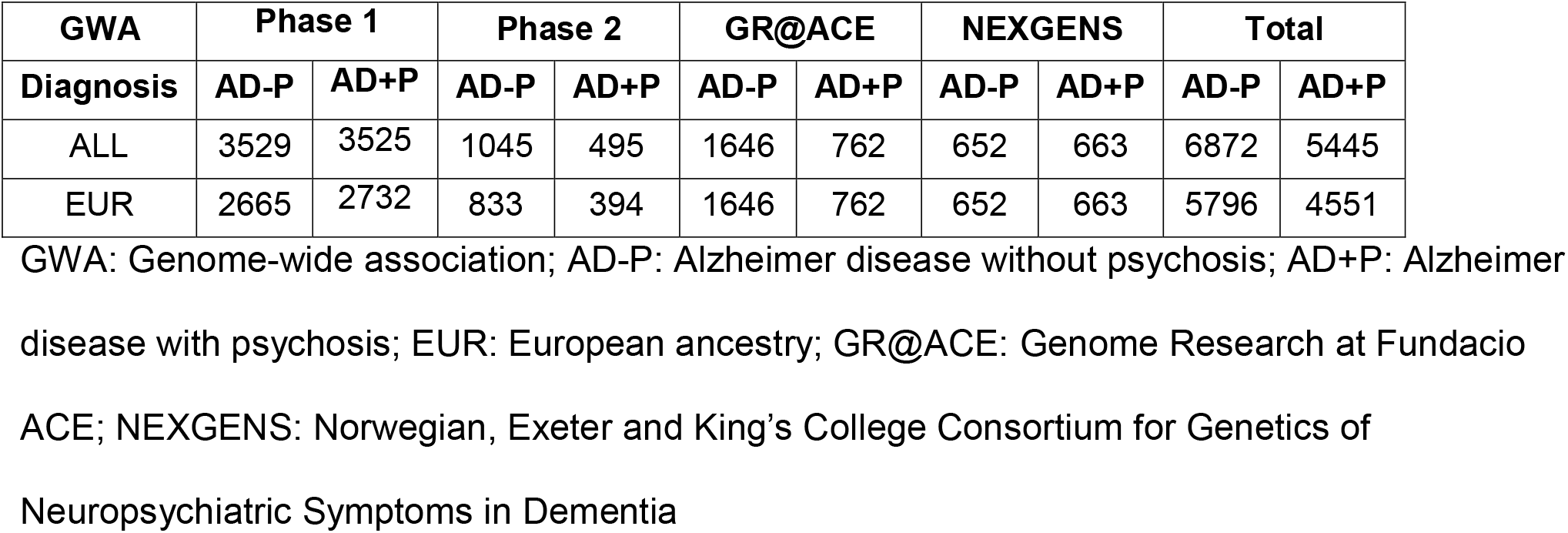
Sample size for each cohort contributing to the meta-analysis.

Genotypes were imputed using the Sanger Imputation Server,^61^ the 1000 Genomes Phase3 reference panel,^62^ and EAGLE2 for pre-phasing^63^ for Phase 1, Phase 2, and GR@ACE. Before imputation, the genotypes were harmonized using the perl script HRC-1000G-check-bim-v4.2.5.pl. This resulted in 85,057,462 imputed or genotyped SNPs for each sample. QC of the imputed SNPs included the requirement that the INFO score for a SNP in each data set > 0.81; MAF > 0.01; and, among all European ancestry subpopulations defined by GemTools, Fst < 0.005. For NEXGENS phasing and imputation was done via the Sanger Imputation Service using the Haplotype Reference Consortium (r1.1) reference panel on all cohorts. After imputation, only SNPs with an imputation quality (INFO) score > 0.4 and MAF > 0.05 were retained.

Separate GWA analyses were performed for the Phase 1, Phase 2, and GR@ACE cohorts, to contrast AD+P versus AD-P for the 9,200,578 SNPs using the Plink option --logistic and with adjustment for the three ancestry dimensions (**Supplementary Figures S3-S5**). For chromosome X, an additional covariate for sex was included. For NEXGENS, separate logistic regressions, implemented in PLINK for each of the five NEXGENS consortium datasets (**Tables S11.1-S11.5**), was used to contrast AD+P versus AD-P for each SNP, with adjustment for the first 10 ancestry principal components. METAL software was used to conduct inverse-variance weighted fixed effects meta-analysis across the five NEXGENS datasets, applying genomic control,^55^ to generate the summary statistics used in the current analysis. The four GWAS statistics (Phase 1, Phase 2, GR@ACE, NEXGENS summary), per SNP, were then meta-analyzed using METAL.

Heritability of AD+P using GenomicSEM was estimated from 1,126,265 summary statistics from our METAL analysis. Of the 7,105,229 SNPs used for GWAS, 1,126,265 matched to those available on the GenomicSEM website. Also, using genome-wide complex trait analysis (GCTA),^64^ heritability was estimated from 9,031 subjects of European ancestry drawn from the Phase 1, Phase 2, and GR@ACE cohorts for which individual genotypes were available (**Table 1B**). Two eigenvectors were used to control for ancestry, 997,105 SNPs were included in the analysis.

Individuals of European ancestry from all four cohorts were used to estimate genetic correlations using LD Score^65^ and LD Hub (version 1.9.3).^66^ We selected phenotypes for analysis based on prior studies showing correlations with psychosis in AD (years of schooling, depressive symptoms) or genetic association with AD+P (schizophrenia), or because they are closely related with the above phenotypes. Specifically, we included intelligence, which is genetically correlated with years of schooling and bipolar disorder which is strongly genetically correlated with both depressive symptoms and schizophrenia. Finally, we included AD as it is a necessary condition of AD±P, and two other neurodegenerative diseases, amyotrophic lateral sclerosis (ALS) and Parkinson’s disease, each of which is associated with a neuropathology that may contribute to psychosis risk in AD.

We evaluated how well three different polygenic risk scores could differentiate 9,031 AD+P and AD-P subjects of European ancestry. We used the pruning and thresholding approach^67^ to compute a PRS for our subjects, developed from GWAS results for AD (PRS_AD_), ^42^ schizophrenia (PRS_SZ_),^68^ and bipolar disorder (PRS_BP_),^69^ separately. We used a set of GWAS p-value thresholds for SNP inclusion in each score (5×10^−8^, 0.0001, 0.001, 0.01, 0.1, 0.2, 0.3, 0.4, 0.5).

Gene-based analyses were performed on the summary association statistics using the most recent version (1.08b) of MAGMA.^70^ For the primary analysis, SNPs were assigned to genes if they lay within the gene boundaries (as defined by NCBI) and the MAGMA “mean” method was used to derive the gene-wide association statistic (the sum of the squared Z statistics for individual SNPs). A secondary analysis assigned SNPs to genes if they lay within 35kb upstream or 10kb downstream of the gene boundary, to capture regulatory regions.^71^

Gene set enrichment analyses were performed in MAGMA,^70^ correcting for the number of SNPs in each gene, linkage disequilibrium (LD) between SNPs and LD between genes. The measure of pathway enrichment is the MAGMA “competitive” test (where the association statistic for genes in the pathway is compared to those of all other protein-coding genes).^72^

Transcriptome-wide association (TWAS) was implemented using the FUSION package^73^ was used to perform a TWAS using dorsolateral prefrontal cortex expression data from the CommonMind Consortium and expression data from 13 Brain tissues from the GTEx (Genotype-Tissue expression) consortium (v7).^74^ Results were corrected for multiple testing of multiple genes within each tissue using the Bonferroni method.

See **Supplementary Methods** for additional details of QC, PRS calculation, pathway analyses and TWAS.

## Results

### Association Analyses

A total of 12,317 subjects, 6,872 AD-P and 5,445 AD+P, were included in this GWAS analysis (**Table 1A**). Contrasting AD-P to AD+P genotypes across the genome revealed two significant loci (**Figure 1, Supplementary Table S1**). One locus was at 4q24, mapping to an intron of *ENPP6* (best SNP rs9994623, O.R. (95%CI) 1.16 (1.10, 1.22), p=1.26×10^−8^). The other locus was at 3p26.1 (best SNP rs201109606, O.R. 0.65 (0.56-0.76), p=3.24×10^−8^). This locus spans the 3’ untranslated region (3’-UTR) of an alternatively spliced variant of *SUMF1* (SUMF1-204 ENST00000448413.5). None of the SNPs showing significant association in these loci are annotated as expression quantitative trait loci (eQTL) in GTEx. Behavior of the association statistics, as assessed by probability-probability plot (**Supplementary Figure S1**), is consistent with the expectation for such analyses, and the genomic control estimate,^75^ GC=1.03, shows no evidence for confounding by ancestry.

**Figure 1.**
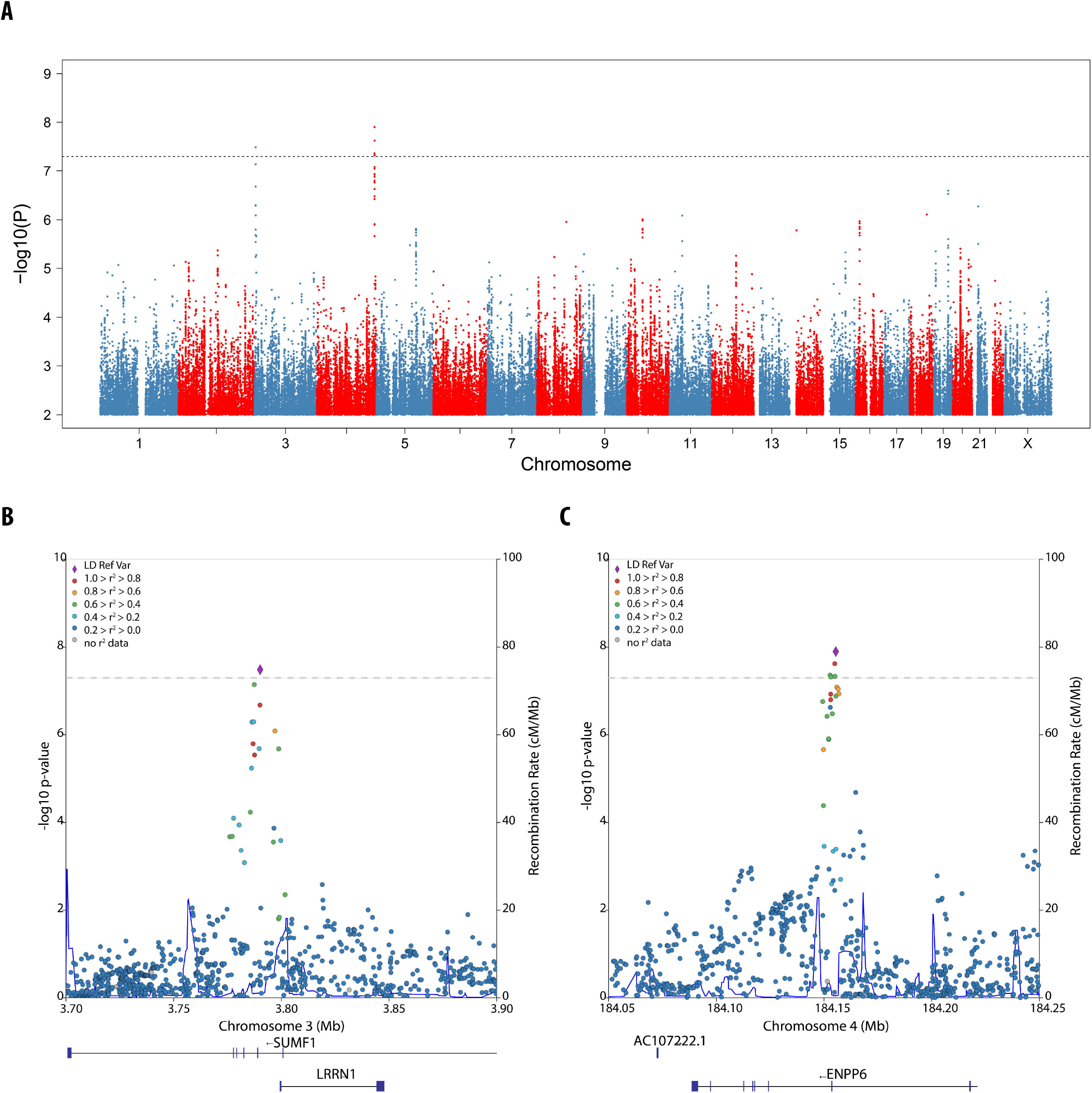
SNP associations with psychosis in AD. **A. Manhattan plot.** The x-axis shows genomic position for autosomes and the X chromosome. The y axis shows statistical significance as −log10 (P). Each point represents an analyzed SNP. The dashed horizontal line represents the threshold for genome-wide significance (p = 5 x 10^−8^). **B-C. Zoom plots of the two genome-wide significant loci**. The x-axis shows genomic position. The left y-axis shows statistical significance as −log10 (P). Each point represents an analyzed SNP, coded by degree of linkage disequilibrium relative to the most significant SNP within the locus. Recombination rate through the region is shown on the right y axis. AD: Alzheimer Disease; LD: linkage disequilibrium; cM: centimorgans; Mb: megabase

For the gene-based tests (**Supplementary Table S2**), only *APOE* (p=1.23×10^−6^) reached the criterion for genome-wide significance (p<2.5×10^−6^).^76^ This association was only significant for SNPs within *APOE* itself. When the 35/10kb window around genes was used to assign SNPs, no genes reached genome-wide significance. There was substantial association signal for SNPs in and near *APOE*, however: the smallest p-value achieved was at rs283811 (z = 5.15, p = 2.55×10^−7^), which falls in an intron of *NECTIN2* (PVRL2 protein). The second smallest p-value occurred for rs429358 (z = 5.12, p = 2.96×10^−7^), which is one of the two SNPs comprising the *APOE* risk haplotype ε4. These two SNPs, separated by 23,441 bp, were in modest LD (r^2^ = 0.52, D’ = 0.91^77^) in the 1000G CEU population sample. To determine if ε4 count could explain the AD+P association signal at this locus, we first analyzed a subset of our subjects who were characterized for the ε4 haplotype (2414 AD+P and 2509 AD-P) by logistic regression of AD+P status (yes/no) on ε4 count, after controlling for three eigenvectors for ancestry. The odds increased significantly with count of ε4 haplotypes (OR = 1.21; 95% CI: 1.11-1.31; p = 8.64×10^−6^). Of the 537 SNPs in this locus, none achieved a p-value<0.0001 in this logistic model after controlling for ε4 count and ancestry (rs2927472 achieved the smallest p-value, 0.00053). Next, using the same subjects, we determined the LD of ε4, in terms of r^2^, with 120 SNPs in the *APOE* locus, all of which had association statistic |z| > 2.0 based on the GWAS of AD+P. We then regressed the statistics for these SNPs, |z|, on their LD with ε4, yielding a strongly positive slope (b=3.14, p = 1.90×10^−41^) and explaining 78.5% of the variance in the observed AD+P z-statistics. Thus, we conclude that the preponderance of AD+P association signal in this locus arises from ε4.

For the pathway enrichment analyses (**Supplementary Table S3**), only the Pathway Interaction Database (PID) IGF1 pathway showed significant enrichment after correction for multiple testing (p=1.17×10^−6^, q=0.011), although it was no longer significant when the 35/10kb window was used (p=0.0469, q=0.920). Interestingly, one of the pathways found to be significantly enriched for AD risk in Kunkle et al.^78^ (GO:48156, tau protein binding) showed significant enrichment (p=6.44×10^−4^ and p=2.21×10^−3^ respectively), albeit not withstanding correction for multiple testing. Given that this pathway includes APOE, the enrichment analysis was repeated excluding genes within 1Mb of APOE (a total of 70 genes), with results shown in **Supplementary Table S3**. Removing the genes in the APOE region greatly reduced the significance of GO:48156 (p=0.0106), suggesting that its enrichment is mainly due to APOE.

TWAS comprised a total of 44,185 gene-tissue combinations (**Supplementary Tables S4 and S5**). No TWAS association was significant after correction for the number of tests performed in all genes and tissues combined (p<1.13×10^−6^, Bonferroni correction for 44,185 tests). Two associations were significant after Bonferroni correction for the number of genes tested in their particular tissue: VN1R108P in GTEx7 hippocampus (p=2.94×10^−6^) and FAM182B in GTEx7 cerebellum (p=5.51×10^−6^). For both genes, an increase in gene expression was associated with the presence of psychosis.

### SNP-Based Heritability

While earlier studies the AD+P phenotype have shown strong clustering in families and substantial heritability, SNP-based heritability has not been estimated. We estimated it in two ways. First, by analyzing our GWAS statistics using GenomicSEM, SNP-based heritability was estimated at 0.181 ± 0.064 (Chi-square = 8.0, df=1, p = .005). An alternative approach, using the GCTA software, evaluated genotypes genome-wide to determine relationships among the samples and how they partitioned within and between AD+P and AD-P sets. This estimate was 0.312 ± 0.053 (Chi-square = 34.98, df=1, p = 3.3×10^−9^). The larger estimate probably arises due to greater information contained in estimated genetic relationships, relative to our modestly powered GWAS, although the heritability estimates are not significantly different. The GCTA analysis focused on subjects of European ancestry, genetically determined, to avoid confounding of ancestry.

### Genetic Correlation, Polygenic Risk Score, and Risk SNP Analyses

Subjects of European ancestry were also used to estimate genetic correlations of AD+P with select phenotypes available from LD Hub (**Table 2**). Consistent with clinical observations, AD+P is significantly genetically correlated with “Years of Schooling” (and nearly so with the related phenotype, “Intelligence”) and with “Depressive Symptoms”. In contrast, AD+P was not significantly genetically correlated with AD (**Table 2**). Nor was AD+P significantly genetically correlated with the two other neurodegenerative disorders evaluated, ALS and Parkinson disease (**Table 2**).

**Table 2.**
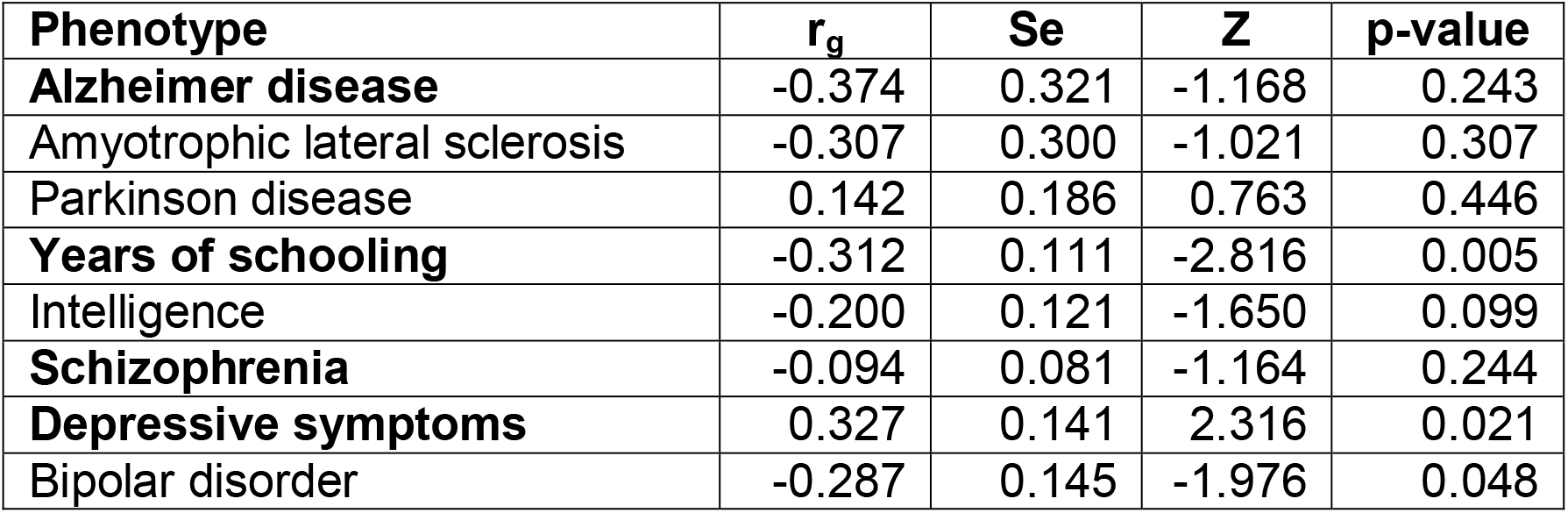
Genetic correlations (r_g_) of psychosis in Alzheimer disease with selected relevant phenotypes. Correlations were obtained from LD Hub.^66^ Phenotypes in bold were chosen, a priori, based on phenotypic or genetic (schizophrenia, bipolar disorder) analyses.

| Phenotype | $r_g$ | Se | Z | p-value |
| --- | --- | --- | --- | --- |
| **Alzheimer disease** | -0.374 | 0.321 | -1.168 | 0.243 |
| Amyotrophic lateral sclerosis | -0.307 | 0.300 | -1.021 | 0.307 |
| Parkinson disease | 0.142 | 0.186 | 0.763 | 0.446 |
| **Years of schooling** | -0.312 | 0.111 | -2.816 | 0.005 |
| Intelligence | -0.200 | 0.121 | -1.650 | 0.099 |
| **Schizophrenia** | -0.094 | 0.081 | -1.164 | 0.244 |
| **Depressive symptoms** | 0.327 | 0.141 | 2.316 | 0.021 |
| Bipolar disorder | -0.287 | 0.145 | -1.976 | 0.048 |

We previously found a significant relationship between risk for AD+P and schizophrenia.^28^ Specifically, we genotyped 94 of 128 SNPs that showed genome-wide-significance for association with schizophrenia in a sample of AD+P subjects. We constructed a predictive score for schizophrenia risk from these SNPs, then assessed whether this score predicted AD+P status in the AD sample. There was a significant negative correlation between the risk score for schizophrenia and AD+P status, which we then replicated by genotyping 60 of the 94 risk SNPs in an independent sample. Now, using SNPs from across the genome and a larger set of AD subjects, results from LD HUB show a negative, but non-significant, genetic correlation with schizophrenia, while showing a negative and significant genetic correlation with bipolar disorder (**Table 2**). In fact, no SNP with p value <10^−4^ for association with psychosis in AD had a p value <10^−5^ in the 108 loci associated with schizophrenia.^79^

Because bipolar disorder and schizophrenia are genetically correlated, we next asked if our original result for the 94 SNPs could be explained by an overlap of risk SNPs for schizophrenia and bipolar disorder. To do so, we tested whether the odds ratios for association of these SNPs for these disorders^69,79^ were independent. They were not (**Supplementary Figure S2**); 91 of 94 SNPs had odds ratios exceeding one for both disorders, whereas 47 were expected under independence (sign test, p = 5.8×10^−20^).

Given the somewhat surprising results for the genetic correlations of AD+P with schizophrenia, bipolar disorder, and AD, we examined whether PRS scores for each of these disorders could differentiate AD+P versus AD-P subjects. In agreement with the genetic correlation, the PRS_BP_ differentiated AD+P from AD-P, whereas PRS_SZ_ showed little ability to differentiate AD+P from AD-P subjects (**Table 3**).

**Table 3.** Prediction of psychosis in Alzheimer disease by polygenic risk scores built using GWAS results for Bipolar disorder, Schizophrenia, and Alzheimer disease, and the pruning and thresholding approach.

**Bipolar Disorder**
| P Value Cut Off | N | OR | I95 OR | u95 OR | P Value | R<sup>2</sup> |
| --- | --- | --- | --- | --- | --- | --- |
| **0.00000005** | 22 | 0.968 | 0.928 | 1.010 | 0.13 | $3.30 \times 10^{-3}$ |
| **0.0001** | 785 | 0.961 | 0.921 | 1.003 | 0.065 | $3.47 \times 10^{-3}$ |
| **0.001** | 3112 | 0.980 | 0.938 | 1.023 | 0.35 | $3.09 \times 10^{-3}$ |
| **0.01** | 14748 | 0.959 | 0.918 | 1.002 | 0.064 | $3.47 \times 10^{-3}$ |
| **0.1** | 79818 | 0.950 | 0.908 | 0.994 | 0.025 | $3.71 \times 10^{-3}$ |
| **0.2** | 134250 | 0.955 | 0.913 | 0.999 | 0.045 | $3.56 \times 10^{-3}$ |
| **0.3** | 182119 | 0.953 | 0.911 | 0.997 | 0.035 | $3.62 \times 10^{-3}$ |
| **0.4** | 225630 | 0.957 | 0.915 | 1.001 | 0.056 | $3.51 \times 10^{-3}$ |
| **0.5** | 265136 | 0.958 | 0.916 | 1.002 | 0.060 | $3.49 \times 10^{-3}$ |

**Schizophrenia**
| P Value Cut Off | N | OR | I95.OR | u95.OR | P Value | R<sup>2</sup> |
| --- | --- | --- | --- | --- | --- | --- |
| **0.00000005** | 329 | 1.051 | 1.008 | 1.096 | 0.021 | $8.02 \times 10^{-4}$ |
| **0.0001** | 3161 | 1.012 | 0.970 | 1.056 | 0.57 | $4.73 \times 10^{-5}$ |
| **0.001** | 8488 | 1.009 | 0.967 | 1.052 | 0.69 | $2.36 \times 10^{-5}$ |
| **0.01** | 26064 | 1.004 | 0.962 | 1.048 | 0.84 | $5.80 \times 10^{-6}$ |
| **0.1** | 99775 | 1.006 | 0.963 | 1.052 | 0.78 | $1.14 \times 10^{-5}$ |
| **0.2** | 153878 | 1.007 | 0.963 | 1.053 | 0.76 | $1.42 \times 10^{-5}$ |
| **0.3** | 198913 | 1.009 | 0.965 | 1.055 | 0.70 | $2.24 \times 10^{-5}$ |
| **0.4** | 238902 | 1.011 | 0.967 | 1.057 | 0.62 | $3.60 \times 10^{-5}$ |
| **0.5** | 274264 | 1.013 | 0.969 | 1.059 | 0.57 | $4.88 \times 10^{-5}$ |

**Alzheimer Disease**
| P Value Cut Off | N | OR | I95.OR | u95.OR | P Value | R<sup>2</sup> |
| --- | --- | --- | --- | --- | --- | --- |
| **0.00000005** | 63 | 1.088 | 1.043 | 1.135 | $9.75 \times 10^{-5}$ | $2.28 \times 10^{-3}$ |
| **0.0001** | 488 | 1.048 | 1.005 | 1.093 | $2.85 \times 10^{-2}$ | $7.18 \times 10^{-4}$ |
| **0.001** | 1962 | 1.059 | 1.015 | 1.105 | $7.76 \times 10^{-3}$ | $1.06 \times 10^{-3}$ |
| **0.01** | 12301 | 1.069 | 1.025 | 1.115 | $1.99 \times 10^{-3}$ | $1.43 \times 10^{-3}$ |
| **0.1** | 83722 | 1.095 | 1.050 | 1.142 | $2.30 \times 10^{-5}$ | $2.69 \times 10^{-3}$ |
| **0.2** | 144621 | 1.096 | 1.051 | 1.143 | $2.12 \times 10^{-5}$ | $2.71 \times 10^{-3}$ |
| **0.3** | 195758 | 1.096 | 1.051 | 1.143 | $2.07 \times 10^{-5}$ | $2.72 \times 10^{-3}$ |
| **0.4** | 239694 | 1.088 | 1.043 | 1.134 | $9.39 \times 10^{-5}$ | $2.29 \times 10^{-3}$ |
| **0.5** | 277645 | 1.088 | 1.043 | 1.135 | $8.80 \times 10^{-5}$ | $2.30 \times 10^{-3}$ |

P Value Cut Off: GWAS p-value threshold used for SNP included in calculating the PRS; N: number of SNPs meeting the threshold; OR: Odds ratio; l95 OR: lower limit of the 95% confidence interval for OR; u95 OR: upper limit of the 95% confidence interval for OR; $R^2$ : partial pseudo- $R^2$ attributable to the PRS after adjusting for the first 2 ancestry eigenvectors.

By contrast, the PRS_AD_ did not agree with the genetic correlation of AD and AD+P from LD Hub. PRS_AD_ significantly predicted AD+P status, in the direction of increased risk for AD+P (**Table 3**). Even when we removed the SNP representing the APOE locus, predictions remained positive (**Supplementary Table 20**). Yet the genetic correlation between our AD+P GWAS and Alzheimer’s disease, as estimated in LD HUB, was negative, although non-significant. Notably, PRS_AD_ was built on results from a larger AD GWAS^42^ than LD Hub uses.^45^ Thus, we conjectured perhaps sample size explains the difference. However, when we computed PRS_AD_ using the GWAS data from^45^, it predicts AD+P as well as PRS_AD_ from the larger GWAS, if not better (cf **Supplementary Table 21** with **Table 3**) Notably, when we analyzed the genetic correlation of the two AD studies in LD Hub, the genetic correlation was 0.9 with standard error of 0.11. Thus, it seems likely that the difference between the genetic correlation estimated by LD HUB and the results for PRS_AD_ trace to the genetic architecture of AD and differences in power for each method.

Because “uncorrelated” is not the same as “independent”, we evaluated one more dimension of these data. Specifically, we evaluated the GWAS-significant SNPs (GWAS SNPs) for schizophrenia,^68^ bipolar disorder,^69^ and AD^42^ to determine whether they also had signal in our AD+P GWAS. We approached this question in two ways. First, we queried the GWAS SNPs to determine if their p-values for AD+P were less than 10^−4^: Eleven SNPs crossed the threshold, all in the *APOE* locus and all associated with AD. Next, we reasoned that if some GWAS SNPs also generated risk for or protection from AD+P, whereas others did not, then those AD+P statistics would be represented by a mixture of distributions. We found support for a mixture of distributions for schizophrenia and separately for AD (**Supplementary Figures S6-S8**), while for bipolar disorder there were too few independent GWAS SNPs to have any confidence in our results (**See Supplementary Methods for details**). Curiously, while AD GWAS SNPs were consistent in their effects on AD and AD+P, effects of schizophrenia GWAS SNPs were not. Instead, risk alleles for schizophrenia could impart risk or protection for AD+P (**Supplementary Methods and Supplementary Figure S9**). In addition, using the mixture model results, we identified a small set of SNPs likely to affect risk to AD+P and either AD or schizophrenia (**Supplementary Table S6**).

## Discussion

We identified evidence of genome-wide significant association with psychosis risk in AD at SNPs within *ENPP6*, and in the 3’-UTR of an alternatively spliced transcript of *SUMF1*. Exploration of multiple data sets did not reveal any current evidence linking the SNPs at these loci to variation in expression of *ENPP6, SUMF1*, or other genes. Similarly, although the alternatively spliced SUMF1-204 transcript is expressed in brain,^80^ AD+P risk SNPs in the SUMF1 locus were not associated with brain expression of SUMF1-204 (S. Sieberts, Personal Communication). Nor were SNPs at these loci linked to other potential genetic mechanisms, such as variation in epigenetic modifications. However, we note that for *SUMF1*, the locus is in the 3’-UTR, a region that often serves a substantial role in regulating protein levels via post-transcriptional mechanisms.^81^

*ENPP6* encodes a glycerophosphodiesterase that is highly expressed in new oligodendrocytes as they differentiate from their precursors.^82^ Recent data in mice have demonstrated that differentiation of oligodendrocytes from their precursors (as indicated by increased *ENPP6* mRNA expression) is a necessary component of early,^83^ i.e. synaptic,^84^ phases of new (motor) learning. ENPP6 protein can be expressed both on the myelin membrane and as a soluble form that is found extracellularly.^85,86^ ENPP6 acts as a hydrolase that severs choline from substrates, including lysophosphatidylcholine, glycerophosphorylcholine, and sphingosylphosphorylcholine.^86^ Of these, it has highest catalytic efficiency towards sphingosylphosphorylcholine,^85^ releasing both sphingosine and phosphocholine. Sphingosine is phosphorylated to generate sphingosine-1-phosphate, which signals via the g-protein-coupled sphingosine-1-phosphate receptor (S1PR1). It is of some interest, therefore, that the S1PR1 modulator, fingolimod,^87^ has been previously shown to increase excitatory synaptic transmission^88^ and improve psychosis-associated behaviors in a genetic animal model of β-amyloid overproduction.^89^

The locus on chromosome 3 maps to introns spanning the 3’-UTR of an alternatively spliced transcript of *SUMF1. SUMF1* encodes formylglycine-generating enzyme, which serves as a master activator of lysosomal sulfatases by converting conserved cysteines to formylglycine in their active sites. As a consequence, genetic disruption of *SUMF1* leads to a multiple sulfatase deficiency syndrome.^90,91^ Importantly, the transcript of *SUMF1* (SUMF1-204, ENST00000448413.5), within which our locus is located, encodes an isoform of formylglycine-generating enzyme (isoform 3, Uniprot Accession Q8NBK3-3) lacking the enzymatically active Cys341 residue.^92^ The functional consequences of this change are not established, but would be anticipated to reduce or eliminate the primary enzymatic function. The function of the novel sequence that replaces the c-terminal of formylglycine-generating enzyme in isoform 3 is also not known, and BLAST of this sequence against the UNIPROT database does not identify homologous proteins. Nevertheless, speaking to the potential functional impact in AD+P, ENST00000448413.5 is detectable in cerebral cortex.^80^

Recently, an appreciation of how lysosomal storage dysfunction also leads to impaired autophagy has emerged.^93^ It is thus not surprising, therefore, that selective depletion of SUMF1 in either astrocytes or neurons results in neurodegeneration.^94^ How alterations in function of formylglycine-generating enzyme, due to a potential change in levels of isoform 3, may modify the course of AD through these mechanisms, and thus result in the AD+P phenotype, remains speculative. We have previously shown, however, that preservation of synaptic protein levels in the context of AD neuropathology is associated with reduced psychosis risk.^89^ Thus, genetic alterations that impact degradation of synaptic proteins by the lysosome to autophagosome pathway are likely to influence risk of psychosis.

We and others^34^ have previously evaluated the association of psychosis in AD with *APOE* risk haplotype ε4, finding inconsistent evidence of association.^34^ Our current findings, obtained from by far the largest cohort to address this question, shed further light on these prior observations. We found that SNPs within *APOE* demonstrate gene-based significant association with AD+P, and that this association appears attributable to the presence of the ε4 *haplotype* itself, without a detectable contribution from other SNPs. Because the impact of ε4 on AD+P risk is not large, increasing the odds by 1.21, prior inconsistent associations with AD+P likely resulted from the much smaller sample sizes in all prior studies.

*APOE* ε4 has been shown to increase the accumulation of amyloid β and phosphorylated tau, and, even in the absence of Aβ overproduction, lead to reductions in dendritic markers and synaptic proteins.^95^ For example, we have shown that human ε4 carriers and mice with targeted replacement of ε4 had down-regulation of numerous glutamate signaling and synaptic proteins.^96^ Increased phosphotau and reduced synaptic proteins (but not altered amyloid β accumulation) have all been associated with psychosis in AD.^97^ Future human and animal model studies of ε4 that control for the contribution of other loci to the genetic risk for AD+P would be helpful in determining the relative contributions of these mechanisms to AD+P.

We previously identified, and independently replicated, an inverse association between polygenic risk for schizophrenia, defined by a limited set of schizophrenia risk SNPs^79^, and risk for psychosis in AD.^28^ It was thus somewhat surprising that we saw a non-significant genetic correlation between these two disorders when considering both a larger set of SNPs and a substantially enlarged cohort of AD subjects with and without psychosis. Instead, we identified a negative genetic correlation with risk for bipolar disorder, a disorder that has substantial genetic overlap with schizophrenia. Because our prior analyses relied on a subset of SNPs significantly associated with risk for schizophrenia, and this set also shows enrichment for affecting risk for bipolar disorder (**Supplementary Figure S2**), this overlap probably explains the discrepancy we now observe between our earlier results and the current results for genetic correlations. However, the lack of genetic correlation of AD+P with schizophrenia conceals an underlying complexity. We observed that schizophrenia risk SNPs evidenced significant mixture regarding AD+P risk, such that risk alleles for schizophrenia could impart risk or protection for AD+P.

In contrast, we observed a positive correlation between genetic risk for depressive symptoms and AD+P, consistent with clinical observations of co-occurrence of depressive and psychotic symptoms in AD patients,^5,13-15^ and evidence that antidepressant medications may have some effect in reducing psychotic symptoms in AD.^98,99^ We also observed a significant negative genetic correlation of educational level with psychosis risk in AD, and a similar pattern, but not quite significant relationship with intelligence. Greater cognitive impairment increases the risk for psychosis in AD; moreover, psychosis in AD is further associated with a more rapid rate of cognitive decline^2-9^ (see also review in^100^). The current findings extend these earlier observations, by showing genetic overlap with measures that may be better construed as indicative of cognitive reserve, as they reflect early life cognitive attainment. Cognitive reserve has long been recognized as protective against developing a degree of cognitive and functional impairment sufficient to lead to a diagnosis of AD.^101^ However, somewhat counter-intuitively, once AD is diagnosed, individuals with greater cognitive reserve decline more rapidly.^101^ Thus, the genetic correlations we observed may point to a biology underlying the presence of greater cognitive impairment in AD, but not the more rapid decline associated with AD+P.

The above findings are subject to several potential limitations. Although our analysis is the largest GWA study of AD+P to date, it is nevertheless modest in sample size in comparison to studies of related complex traits.^78,79^ As our heritability results show, a substantial increase in sample size will identify many additional loci as having a significant association with psychosis risk in AD. Similarly, increased sample size is needed to provide the necessary power to identify genes, transcripts, genetically correlated traits, and pathways in the corresponding analyses that derive from the SNP-based associations.

One of our GWAS-significant SNPs, rs201109606, falls in a genomic region marked as simple repeats. Perhaps because the SNP is difficult to impute, it did not pass QC for some of our data. Nonetheless, the largest two of the four datasets contribute to this result and the estimates of the odds ratios for the two data sets are remarkably similar, 0.684 and 0.637. Moreover, other SNPs at this locus also support the findings (**Supplementary Table S1**), although those associations are not quite GWAS significant. Thus, this result requires replication. For additional discussion of other findings and potential limitations, see **Supplementary Material**.

Currently established treatments for psychosis in AD patients are suboptimal, perhaps reflecting in part that these treatments were not derived to prevent or reverse an identified biology of AD+P.^100^ The development of effective, specific, therapeutic targets will therefore require as a first step delineating this underlying biology. Our study provides the first unbiased evidence of association of specific genetic loci with psychosis in AD and can thus serve as an initial road map to AD+P biology. These findings, in conjunction with available functional genomic and post-mortem data, provide multiple links to mechanisms influencing synaptic function as contributors to psychosis in AD.

## Data Availability

Data will be submitted to NIAGADS (https://www.niagads.org) upon acceptance of peer-reviewed manuscript.

## Acknowledgments

This study was supported by the following federal grants: AG027224 (RAS), MH116046 (RAS), MH057881 (BD), AG030653 (MIK), AG041718 (MIK), AG066468 (OLL)

A complete acknowledgements list of contributing individuals, consortia, and their grant support can be found in Supplementary Material.

## Author Contributions

Each author is expected to have made substantial contributions:

1. to the conception **or** design of the work; RAS, BD, MIK, CB, OLL
2. to the acquisition, analysis, **or** interpretation of data; MAAD-S, LK, BC, JCH, EAW, LM, RS, IH, SM-G, LT, MB, EA-M, SV, YL, BH, DA, GS, SB, AR, IS, HKS, BE, ES, OAA, SD, LA, DS, BB, DA, GF, PM, AS, DDR, AP, JW, RM, TF, AR, CB, PH, OLL, MIK, BD, RAS
3. to the creation of new software used in the work; YL
4. have drafted the manuscript or substantively revised it. MAAD-S, JCH, LK, BC, LM, CB, PH, MIK, BD, RAS

Each author must have approved the submitted version (and any substantially modified version that involves the author’s contribution to the study) AND to have agreed both to be personally accountable for the author’s own contributions and to ensure that questions related to the accuracy or integrity of any part of the work, even ones in which the author was not personally involved, are appropriately investigated, resolved, and the resolution documented in the literature.

## URLs

METAL http://csg.sph.umich.edu/abecasis/metal/index.html

GemTools http://www.compgen.pitt.edu/GemTools/GemTools.htm

PLINK https://www.cog-genomics.org/plink2/

NCBI RS names https://ftp.ncbi.nih.gov/snp/redesign/latest_release/VCF/

LD-Hub: http://ldsc.broadinstitute.org/LDHub/

Sanger Imputation Service: https://imputation.sanger.ac.uk

Psychiatric Genetics Consortium: https://www.med.unc.edu/pgc/

Imputation preparation: https://www.well.ox.ac.uk/∼wrayner/tools/

CMC/AMP-AD eQTL Meta-analysis: https://www.synapse.org/#!Synapse:syn16984815

NCBI gene2go file: ftp://ftp.ncbi.nlm.nih.gov/gene/DATA/

Reactome: https://reactome.org/download-data

Gene Ontology: http://geneontology.org/docs/download-ontology/

Molecular Signatures Database: https://www.gsea-msigdb.org/gsea/msigdb/index.jsp

R package qvalue: http://github.com/jdstorey/qvalue

LD Score Regression (LDSC) software v1.0.0 : https://github.com/bulik/ldsc

HapMap 3 https://www.sanger.ac.uk/resources/downloads/human/hapmap3.html

Common mind consortium: https://www.nimhgenetics.org/resources/commonmind

FUSION software http://gusevlab.org/projects/fusion/

